# Comparing the effects of a short nap and non-sleep deep rest on perceptual, cognitive, and physical performance in active adults

**DOI:** 10.64898/2026.03.03.26347495

**Authors:** Omar Boukhris, Haresh Suppiah, Matthew Driller

## Abstract

This study compared the effects of a 25-min nap opportunity and a 10-min non-sleep deep rest (NSDR) condition on perceptual, cognitive, and physical performance in physically active young adults. Sixty participants (26 female, 34 male; 22 ± 4 years) were randomly assigned to one of three groups (nap, NSDR, control; n = 20 each). All groups completed identical assessments immediately, 20 min, and 40 min post-intervention. Mixed-effects models, adjusted for sex, prior-night sleep, and weekly physical activity, revealed a significant Group × Time interaction for sleepiness, fatigue, readiness to perform, and handgrip strength (p < 0.05). At 40 min post-intervention, the nap group reported lower fatigue than control and higher readiness to perform than both control and NSDR (p < 0.05). No significant effects were observed for the NSDR condition on perceptual, cognitive, or physical outcomes (p > 0.05). These findings indicate that a short nap can enhance perceived readiness and reduce fatigue after a brief latency period, whereas NSDR did not elicit significant effects under the present conditions.

## Introduction

Insufficient sleep and poor sleep quality are common across students, professionals, and athletes, often driven by busy schedules, competition demands, academic pressures, travel, and stress (Gardani et al., 2022; Walsh et al., 2021). These disturbances impair attention, memory, emotional regulation, and daily performance (Metschura, 2023), and excessive daytime sleepiness has been linked to poorer academic outcomes (Nguyen et al., 2025). Comparable effects are observed in workplace and sport settings, where inadequate sleep reduces performance, disrupts mood, elevates stress, and delays recovery (Driller et al., 2023; Pilcher & Morris, 2020). To mitigate these effects, various techniques have been explored, including daytime napping (Boukhris, Trabelsi, et al., 2024; Boukhris et al., 2025), mindfulness and meditation (Jones et al., 2020; Li et al., 2018), and biofeedback-assisted breathing exercises (Li et al., 2022).

Recently, non-sleep deep rest (NSDR) has emerged as an innovative recovery method involving guided relaxation without the need for sleep or extensive training (Boukhris, Suppiah, et al., 2024). NSDR protocols commonly include a structured sequence of techniques such as body scans (i.e., systematically shifting attention from the toes to the head), slow nasal breathing, and focused awareness on internal sensations or external sounds. NSDR may facilitate a state that differs from sleep itself but appears to offer some of the same restorative benefits (Boukhris, Suppiah, et al., 2024; Wrzeciono et al., 2024). NSDR may enhance performance and mood by promoting parasympathetic dominance, resulting in reduced stress and improved autonomic balance (Boukhris, Suppiah, et al., 2024; Wrzeciono et al., 2024). Additionally, from psychological perspective, guided relaxation and visualization techniques inherent in NSDR may reduce anxiety and enhance mood by facilitating mental detachment and promoting relaxation (Nien et al., 2023). Our previous work in 65 participants demonstrated that a 10-minute NSDR condition significantly enhanced physical performance, cognitive function, and mood-related measures immediately following the intervention (Boukhris, Suppiah, et al., 2024). Despite these promising findings, direct comparisons of NSDR to napping strategies, particularly daytime napping, remain notably absent in the current literature, highlighting a critical knowledge gap. This comparison is essential because NSDR is often described as producing restorative benefits similar to sleep, but through conscious relaxation rather than actual sleep. If NSDR can replicate or even approximate the physiological and perceptual effects of a nap without inducing sleep inertia or requiring a sleep-conducive environment, it could represent a highly practical alternative for individuals with limited time or opportunities to rest. Further, for those individuals who cannot nap during the day, NSDR may be a viable alternative.

Daytime naps are widely recognised for their positive effects on recovery, cognitive function, and physical performance (Boukhris, Trabelsi, et al., 2024; Boukhris et al., 2025; Mesas et al., 2023); however, they may also present practical drawbacks, such as the requirement for a suitable environment, variable nap durations, and sleep inertia, a temporary reduction in performance and alertness following awakening, lasting up to an hour (Mesas et al., 2023). Conversely, NSDR offers potential advantages, including a shorter and more consistent duration (approximately 10 minutes), minimal preparation, absence of sleep inertia, and greater feasibility for individuals with demanding schedules or environments lacking conducive conditions for napping (Boukhris, Suppiah, et al., 2024).

Therefore, the present study aims to directly compare the effectiveness of a brief NSDR condition, a 25-minute nap opportunity, and a control condition on perceptual, cognitive, and physical performance outcomes in physically active individuals. Clarifying whether NSDR yields comparable or superior benefits to napping will provide valuable insights for developing practical strategies to support alertness and performance in physically active individuals.

## Methods

### Participants

Sixty physically active young adults (26 female, 34 male; mean age 22 ± 4 years) participated in the current study. Participants were randomly allocated to one of three conditions: nap, NSDR, or control (n = 20 per group). On average, participants reported engaging in 8 ± 3 hours of physical activity per week across a variety of sports (e.g., Australian football, soccer, rugby, basketball). All participants were free from musculoskeletal injuries and self-reported sleep or medical disorders, and were not using medications known to affect sleep, cognition, or physical performance. Written informed consent was obtained prior to enrolment. The study was conducted in accordance with the Declaration of Helsinki and approved by the local institutional Human Ethics Committee.

### Experimental design

The study employed a randomised, parallel-group design with three experimental arms: a 25-min nap opportunity, a 10-min non-sleep deep rest condition (NSDR), and a no-intervention control. Participants were randomly allocated to one of the three groups, and assessments were conducted at three time points: immediately, 20 min, and 40 min post-intervention (Figure 1). Randomisation was performed by preparing identical papers labelled with each group, which were mixed and distributed to participants. Each participant opened their assigned paper to reveal their group. Participants were instructed to refrain from caffeine, alcohol, and vigorous exercise for at least 4 h before testing. We employed a post-only assessment schedule following all three conditions to characterise the time-course of performance readiness while minimising pre-test contamination. Same-session pre-testing was avoided because it can elevate arousal, induce fatigue, and introduce practice effects that may attenuate intervention effects for relaxation-based protocols (nap/NSDR). The 0–40 min window was chosen to capture recovery from any transient sleep inertia after the 25-min nap and to align with real-world use cases (e.g., return-to-play intervals).

**Figure 1.**
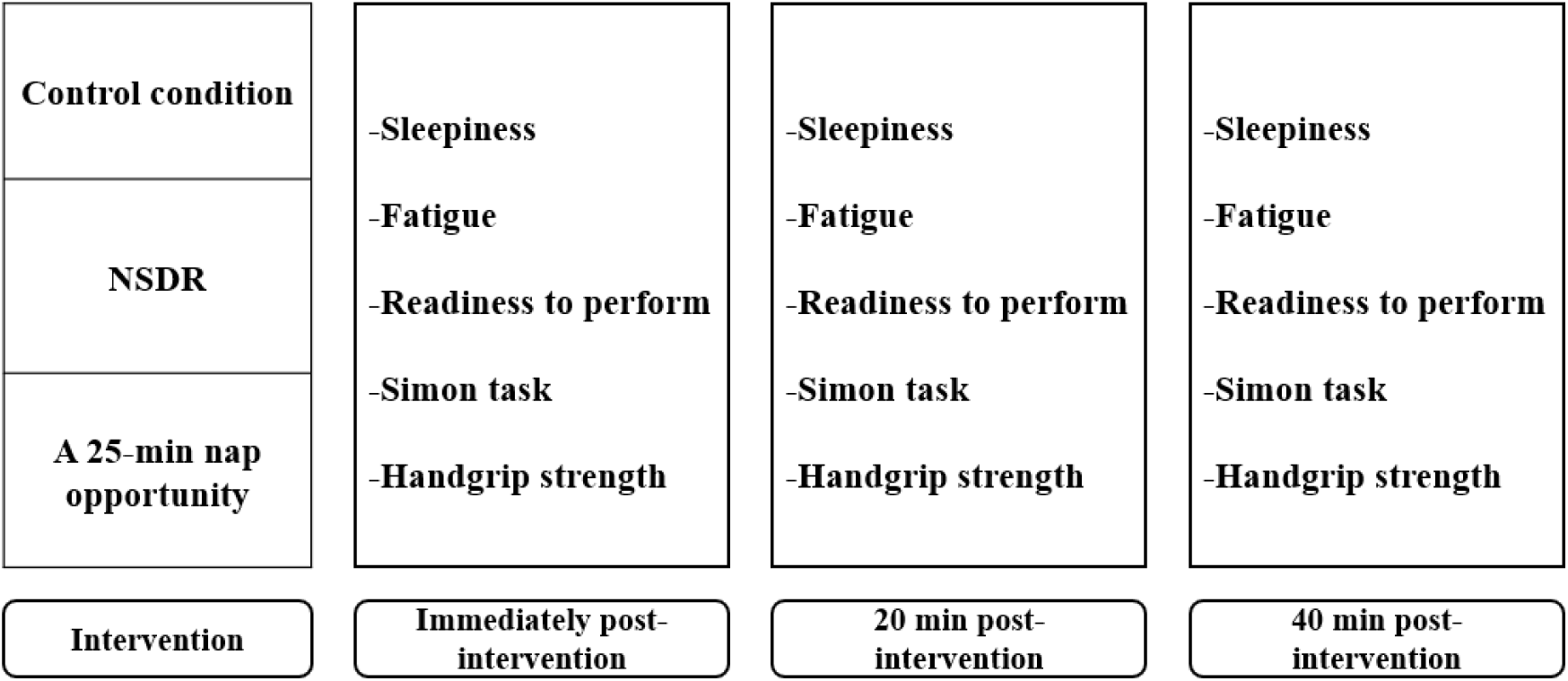
Schematic representation of the experimental design.

For the nap condition, the session was performed in a quiet, dimly lit sleep room (< 5 lux) equipped with a napping pod (Podtime, Restworks, Australia) (Figure 2). The environment was maintained at 21-23 °C to optimise comfort. The nap opportunity took place between 14:35 h and 15:00 h, and sleep was monitored using the Somfit wearable EEG device (Compumedics Ltd., Australia).

**Figure 2.**
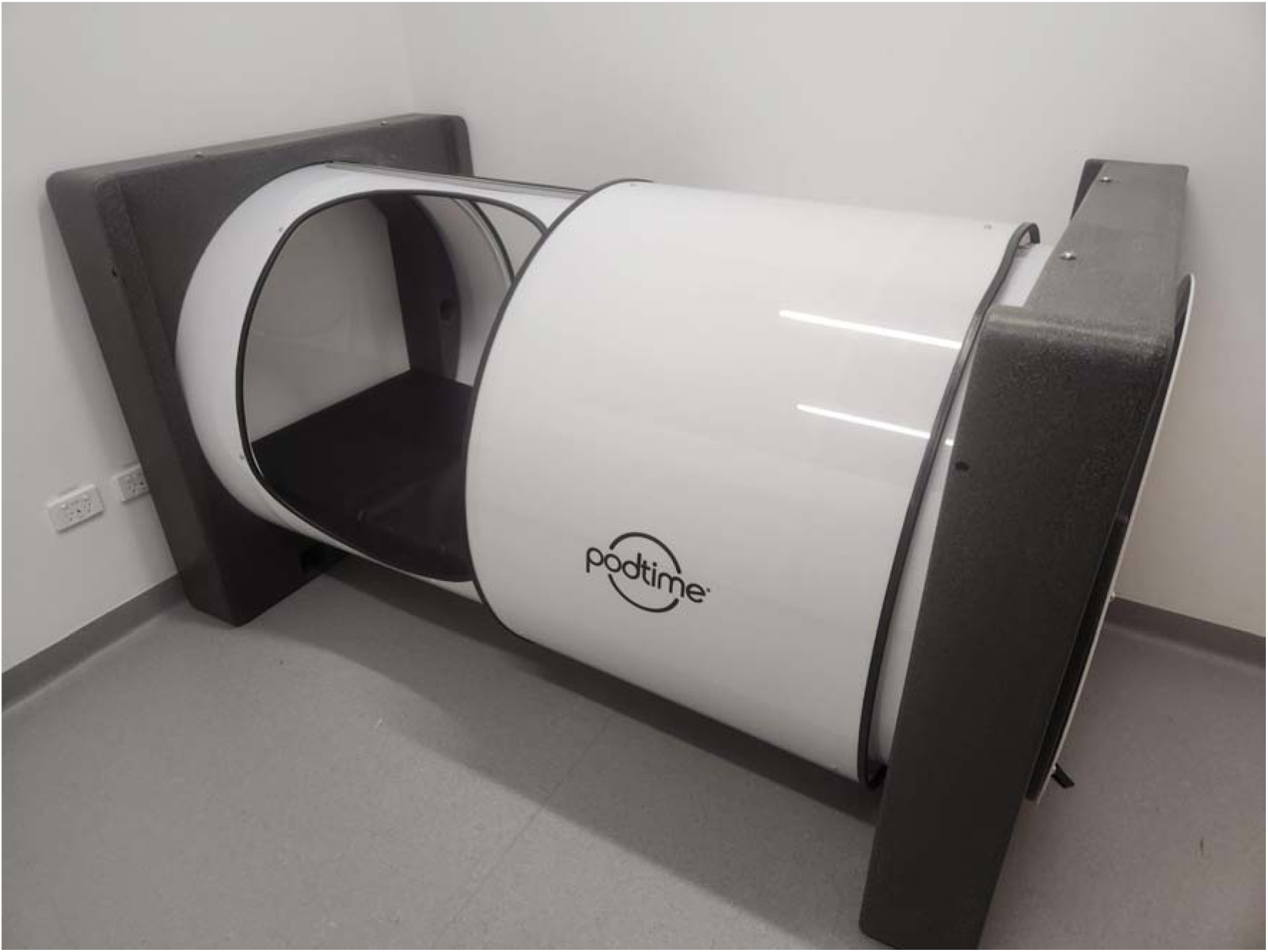
A photo of the napping pod where the 25-minute nap opportunity took place.

For the NSDR and control conditions, testing was conducted at 15:00 h. The NSDR group completed a 10-min guided session delivered through a standardised audio protocol (https://youtu.be/AKGrmY8OSHM), lying comfortably on mats in a quiet, dimly lit room (< 20 lux). In the control condition, participants sat quietly in the same environment under supervision, without engaging in relaxation exercises, sleep, or the use of electronic devices. Physiological responses (heart rate (HR), HR variability (HRV), and skin temperature) were continuously recorded in both NSDR and control using a smart ring (Ultrahuman Ring AIR, Ultrahuman Healthcare, Bangalore, India).

Following the intervention period, participants completed perceptual (sleepiness, fatigue, readiness to perform), cognitive (Simon task), and physical (handgrip strength) assessments at each of the three post-intervention time points.

### Sleep monitoring during naps

Sleep during the nap opportunity was assessed using a wearable EEG device (Somfit, Compumedics, Australia). The Somfit is a portable system that attaches to the forehead with an adhesive patch and transmits data to a companion mobile application (iOS/Android). The Somfit records single-channel EEG (Fp1–Fp2), electro-oculography (Fp1–Fpz, Fp2–Fpz), electromyography (Fp1–Fp2), pulse oximetry, head position, snoring sound, movement, and ambient light. Validation against polysomnography (PSG) has shown an overall agreement of 76.1% between Somfit’s automated scoring and consensus PSG hypnograms, with a Cohen’s kappa of 0.67. Sleep/wake classification accuracy was 89.3%, comparable to inter-scorer variability observed among PSG technologists (McMahon et al., 2024).

### Physiological monitoring during NSDR and control conditions

Physiological responses during the NSDR and control conditions were continuously recorded using a finger-worn smart ring device (Ultrahuman Ring Air, Ultrahuman Healthcare, Bangalore, India). The ring integrates a photoplethysmography (PPG) sensor to capture heart rate (HR) and heart rate variability (HRV, expressed as RMSSD), along with a non-contact sensor to monitor peripheral skin temperature. For each 10-min intervention, start and end values of HR, HRV, and temperature were extracted, as well as average values across the session (for HRV and temperature) and the minimum HR reached during the period.

### Perceptual measures

Subjective sleepiness was assessed using the Stanford Sleepiness Scale, where participants rated their current level of alertness on a 7-point scale ranging from 1 (“active, alert, fully awake”) to 7 (“near sleep onset, struggling to remain awake”). This tool has shown good validity and reliability, with 88% agreement in prior assessments (Hoddes et al., 1973).

Whole-body fatigue was measured on a 10-point numerical scale (Romyn et al., 2022), with 0 representing “normal, without pain or stiffness” and 10 reflecting “extreme fatigue.”

Readiness to perform was evaluated using a comparable 10-point scale (Romyn et al., 2022), in response to the question, “How ready are you to perform?” where 0 indicated “not at all ready” and 10 indicated “extremely ready.”

### Cognitive performance

Cognitive control was assessed using the Simon task, a well-established measure of interference control and response inhibition (Simon, 1990). Participants were seated at a laptop and completed the task through an open-source software platform (The Psychology Experiment Building Language, PEBL, Version 2.1) (Mueller & Piper, 2014). They were instructed to respond as quickly and accurately as possible to the colour of a circle (red or blue) that appeared to the left or right of a central fixation point, while ignoring its spatial location. Responses were made using the thumbs: the left shift key for red and the right shift key for blue.

The task consisted of 70 randomly ordered trials, with equal proportions of congruent trials (stimulus location matched response side) and incongruent trials (stimulus location opposite to response side). Each stimulus remained on screen until a response was given or for a maximum of 1.5 seconds, after which the next trial appeared. Two performance outcomes were calculated: mean reaction time for correct responses, reflecting processing speed, and percentage of correct responses, reflecting accuracy.

### Physical performance

Maximal grip strength of the dominant hand was assessed using a Jamar Plus Digital hand dynamometer (Paterson Medical, Green Bay, WI, USA). The device was individually adjusted to accommodate hand size, and measurements were taken in a standardised position: participants stood upright with shoulders adducted and neutrally rotated, elbows flexed at 90°, forearms in neutral alignment, wrists extended between 0°-30° with 0-15° ulnar deviation, and feet flat on the floor. Each participant performed three maximal-effort trials, separated by 30 seconds of passive rest, and the highest value (kg) was used for analysis. This protocol is considered the gold standard for grip strength assessment and has demonstrated excellent validity and reliability (Hamilton et al., 1994).

### Statistical analysis

All data were analysed using R (version 4.5.1; R Core Team, Vienna, Austria). Linear mixed-effects models (LMMs) were used to examine the effects of Group (Nap, NSDR, Control), Time (immediately post, 20-min post, 40-min post), and their interaction on perceptual (sleepiness, fatigue, readiness to perform), cognitive (reaction time and accuracy during the Simon task), and physical outcomes (handgrip strength). Participant ID was included as a random effect to account for repeated measures. Sex, self-reported sleep duration, and weekly physical activity were entered as covariates in all models.

Model fit and variance components were quantified using intra-class correlation coefficients (ICC), marginal R² (variance explained by fixed effects), and conditional R² (variance explained by fixed + random effects).

When significant interaction effects were detected, pairwise post-hoc comparisons were performed using estimated marginal means with Bonferroni correction to adjust for multiple testing. Effect sizes were expressed as partial eta squared (ηL²) with values interpreted as: small (0.01), medium (0.06), and large (0.14) (Lakens, 2013).

The significance level was set at p < 0.05. Exact p-values are reported; those below 0.001 are denoted as p < 0.001.

## Results

The characteristics of participants within the subgroup samples are summarised in Table 1, while Table 2 presents sleep architecture during the 25-min nap opportunity and physiological measures (HR, HRV, and skin temperature) recorded during the NSDR and control conditions.

**Table 1.** Participant characteristics in the control, non-sleep deep rest (NSDR), and nap groups.

|  | **Control group** | **NSDR group** | **Nap group** |
| --- | --- | --- | --- |
| **Female participants (n)** | 7 | 9 | 10 |
| **Male participants (n)** | 13 | 11 | 10 |
| **Age (years)** | 21 ± 2 | 21 ± 3 | 25 ± 4 |
| **Body Mass (kg)** | 74 ± 11 | 72 ± 14 | 66 ± 10 |
| **Height (cm)** | 176 ± 10 | 173 ± 10 | 168 ± 10 |
| **Physical activity (hours/week)** | 9 ± 3 | 9 ± 3 | 7 ± 2 |
| **Previous night's sleep duration (H:min)** | 7:09 ± 1 | 6:33 ± 1 | 7:10 ± 1 |

**Table 2.**
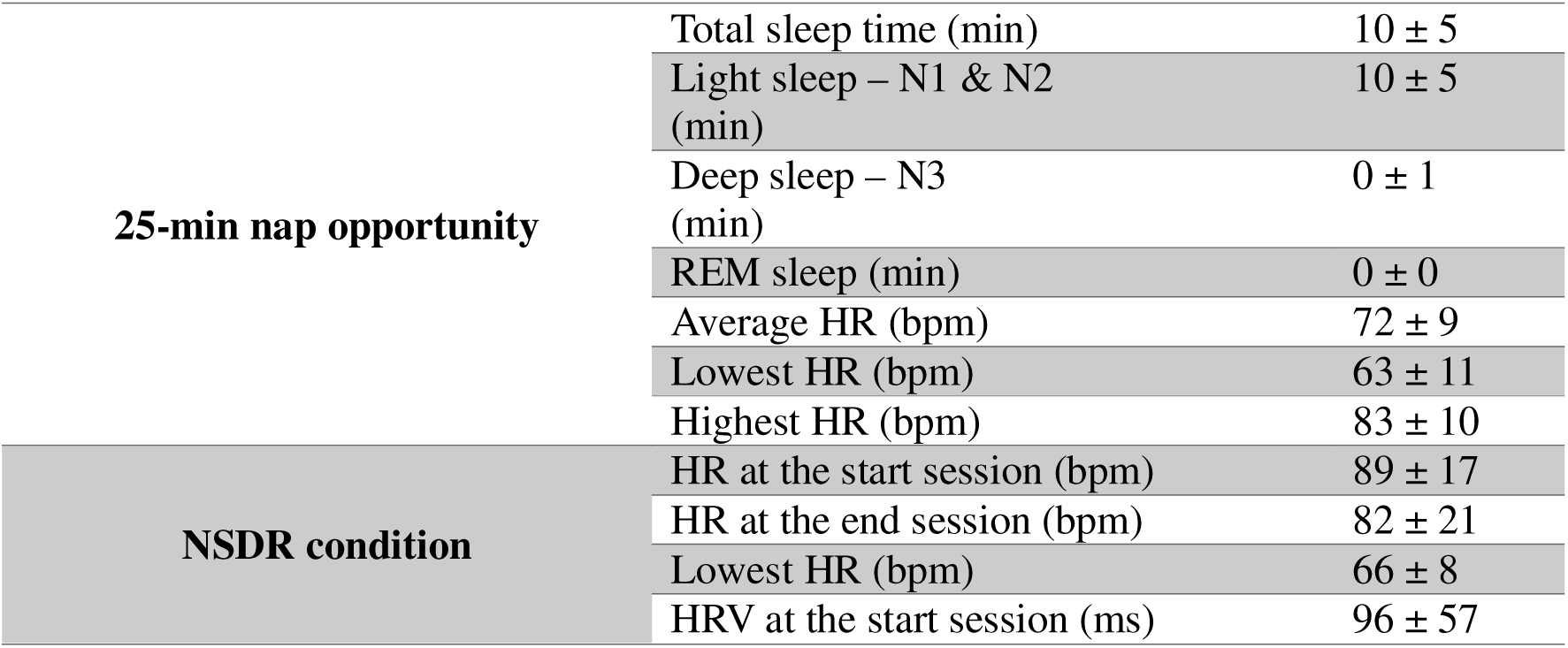
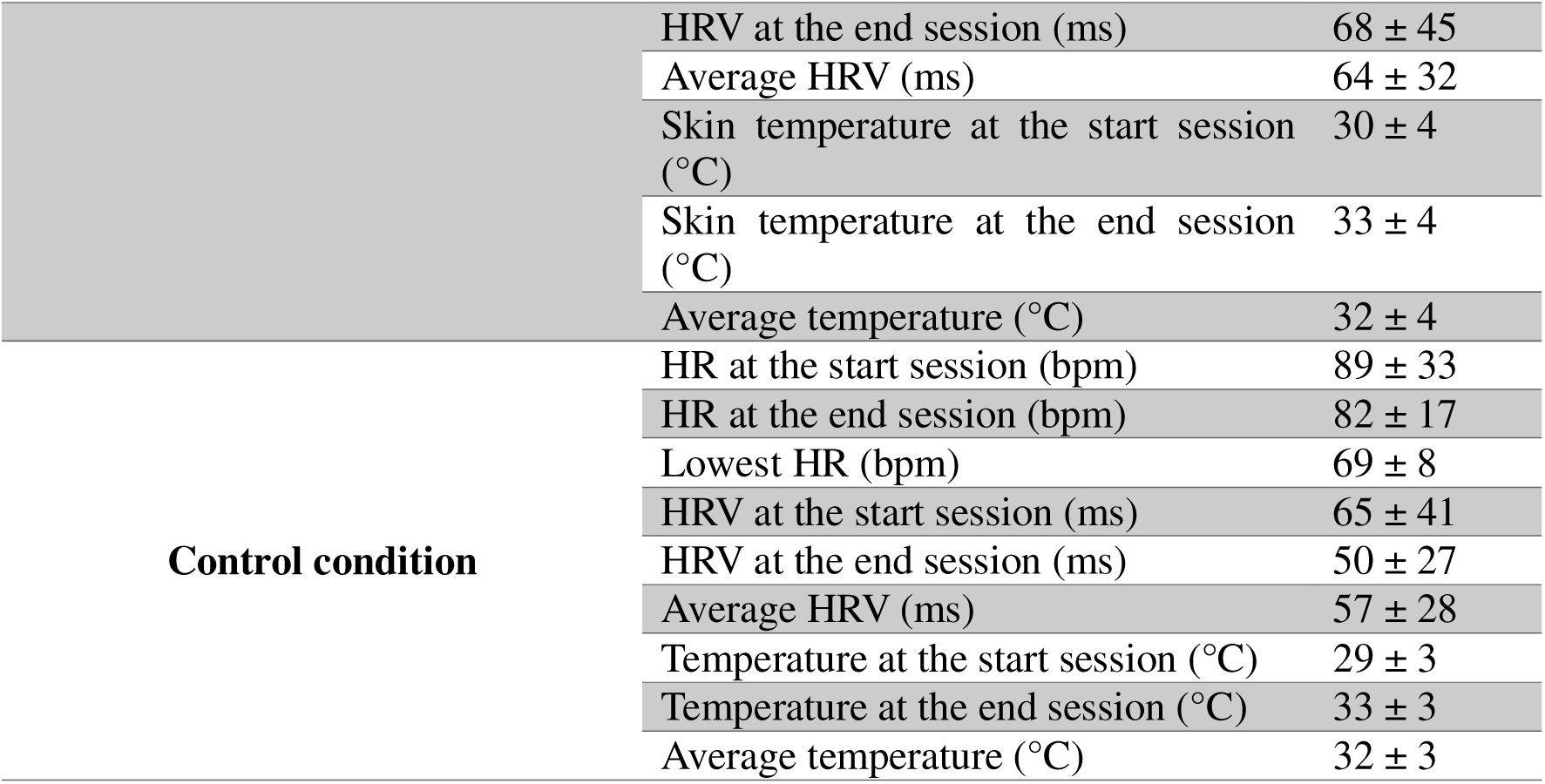
Sleep architecture during the 25-min nap opportunity and physiological measures (heart rate (HR), heart rate variability (HRV), and skin temperature) during NSDR and control conditions.

The linear mixed-effects models are presented in Table S1, and model fit indices (ICC, marginal R², and conditional R²) are reported in Table S2.

### Perceptual measures Sleepiness

There was an interaction between Group and Time for sleepiness (F(_4,109)_ = 4.83, p = 0.001). While groups did not differ at any single timepoint, the nap group showed within-condition reductions from post-intervention to 20 min (p = 0.008) and to 40 min (p < 0.001). The additional drop from 20 to 40 min was small and did not meet the Bonferroni-adjusted threshold (p = 0.04).

### Fatigue

A significant Group × Time interaction was observed for fatigue (F_(4,109)_ = 8.74, p < 0.001). At 40 min post-intervention, the nap (ΔEMM = 1.59, p = 0.01) group reported significantly lower fatigue than the control group. No differences were observed between NSDR and control groups (p > 0.05). Within the nap condition, fatigue was significantly lower at 40 min compared with immediately (p < 0.001) and 20 min (p = 0.02) post-intervention.

### Readiness to perform

A significant Group × Time interaction was observed for readiness to perform (F_(4,112)_ = 9.16, p < 0.001). At 40 min post-intervention, the nap group reported significantly higher readiness to perform than the control (ΔEMM = -2.33, p < 0.001, Cohen’s *d* = 1.48) and NSDR (ΔEMM = 2.21, p = 0.001, Cohen’s *d* = 1.25) groups. No differences were observed between NSDR and control groups (p > 0.05). Within the nap condition, readiness to perform was significantly higher at 20 min (p = 0.01) and 40 min (p < 0.001) compared with immediately post-intervention, and also higher at 40 min compared with 20 min (p = 0.01).

**Figure 3.**
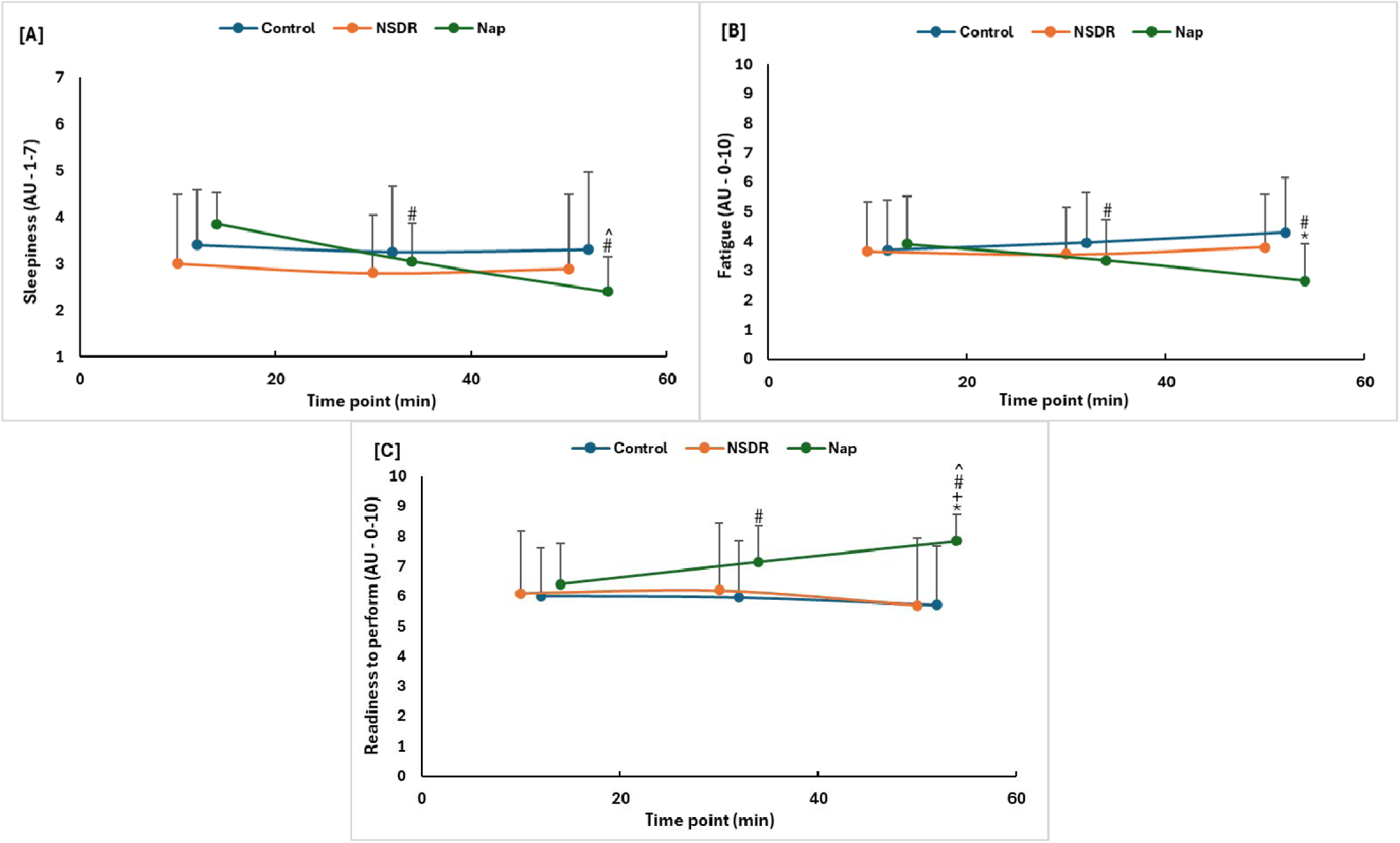
Comparison of sleepiness [A], fatigue [B], and readiness to perform [C] scores between nap, non-sleep deep rest (NSDR) and control groups at post, 20-min, and 40-min assessments. *: significant difference compared with control group; +: significant difference compared with NSDR group; #: significant difference compared with immediately post; ^: significant difference compared with 20-min post.

### Cognitive performance

No significant Group × Time interaction was observed for reaction time (F_(4,107)_ = 2.27, p = 0.06) and accuracy percentage during the Simon task (F_(4,107)_ = 1.25, p = 0.29).

### Physical performance

There was an interaction between Group and Time for handgrip strength (F_(4,113)_ = 4.00, p = 0.004). While groups did not differ at any single timepoint, the NSDR group showed small within-condition reductions from post-intervention to 40 min (p = 0.03).

**Table 3.** Comparison of perceptual, cognitive, physical measures between nap, non-sleep deep rest (NSDR) and control groups at post, 20-min, and 40-min assessments. Bold values represent significant group x time interactions (p < 0.05).

| | Control | | | NSDR | | | Nap | | | Group × Time | |
| --- | --- | --- | --- | --- | --- | --- | --- | --- | --- | --- | --- |
| | Immediately post | 20 min post | 40 min post | Immediately post | 20 min post | 40 min post | Immediately post | 20 min post | 40 min post | P value | Effect size ($\eta^2$) |
| Sleepiness (AU) | 3.4 ± 1.2 | 3.3 ± 1.4 | 3.3 ± 1.7 | 3.0 ± 1.5 | 2.8 ± 1.2 | 2.9 ± 1.6 | 3.9 ± 0.7 | 3.1 ± 0.8 | 2.4 ± 0.8 | **0.001** | 0.15 |
| Fatigue (AU) | 3.7 ± 1.7 | 4.0 ± 1.7 | 4.3 ± 1.9 | 3.7 ± 1.7 | 3.6 ± 1.6 | 3.8 ± 1.8 | 3.9 ± 1.6 | 3.4 ± 1.4 | 2.7 ± 1.3 | **<0.001** | 0.24 |
| Readiness to perform (AU) | 6.0 ± 1.6 | 6.0 ± 1.9 | 5.7 ± 1.9 | 6.1 ± 1.6 | 6.2 ± 2.2 | 5.7 ± 2.3 | 6.4 ± 1.4 | 7.2 ± 1.2 | 7.9 ± 0.9 | **<0.001** | 0.25 |
| Reaction time (ms)<sup>^</sup> | 434 ± 48 | 411 ± 47 | 405 ± 43 | 409 ± 45 | 397 ± 46 | 402 ± 39 | 433 ± 63 | 409 ± 47 | 400 ± 41 | 0.06 | 0.08 |
| Accuracy (%) | 93 ± 3 | 93 ± 4 | 93 ± 4 | 94 ± 4 | 93 ± 5 | 95 ± 5 | 94 ± 4 | 95 ± 3 | 96 ± 2 | 0.29 | 0.04 |
| Handgrip strength (kg) | 44 ± 15 | 42 ± 14 | 42 ± 13 | 43 ± 15 | 41 ± 13 | 40 ± 13 | 36 ± 11 | 37 ± 11 | 38 ± 11 | **0.004** | 0.13 |

<sup>^</sup> reaction time measures taken from correct trials during the Simon task.

## Discussion

This study is, to our knowledge, the first to compare a 25-min nap opportunity with an NSDR protocol in physically active adults. Participants who completed the nap condition reported lower fatigue compared with those in the control condition and greater readiness to perform compared with both the control and NSDR conditions at 40 min post-intervention. These findings suggest that a brief nap may promote higher perceived readiness and lower fatigue in the period following the intervention, although these benefits appear to emerge after a short delay, possibly reflecting the dissipation of sleep inertia. No significant effects were found in favour of NSDR across any of the measured outcomes.

Perceptual responses showed that only the nap condition influenced subjective states. Participants in the nap group reported lower fatigue compared with the control group and greater readiness to perform compared with both the control and NSDR groups at 40 min post-intervention. These effects appeared after a short delay, likely reflecting the gradual dissipation of sleep inertia typically observed within the first hour after waking (Mesas et al., 2023). In contrast, NSDR did not lead to significant changes in any perceptual measures, which differs from our previous study that reported improved fatigue, alertness, and mood following NSDR conditions (Boukhris, Suppiah, et al., 2024). While both interventions encourage relaxation, only the nap involved actual sleep, which likely activated physiological processes that support reductions in fatigue and enhanced readiness. The statistical indices support this interpretation. The high intraclass correlations for fatigue (ICC = 0.75) and readiness to perform (ICC = 0.81) indicate stable individual ranking across repeated measures, while the low marginal R² values (0.09-0.15) show that group allocation explained only a small proportion of the variance. Together, these findings suggest that interindividual factors may have played a stronger role than intervention type in shaping perceptual responses to NSDR, whereas the nap produced distinct significant effects despite this variability.

Cognitive performance, assessed through reaction time and accuracy in the Simon task, did not differ significantly between the nap, NSDR, and control conditions at any time point. These null findings suggest that neither intervention produced measurable effects on processing speed or response accuracy under well-rested conditions. The absence of significant differences aligns with the idea that both interventions primarily influence perceptual or subjective states rather than higher-order cognitive processing when baseline alertness is already sufficient. It is also possible that the short task duration and lack of prior fatigue limited the sensitivity to detect significant changes in executive performance. Another consideration is that cognitive performance tends to show high interindividual stability, making between-group effects more difficult to detect. This is reflected in the present model statistics, where reaction time and accuracy exhibited high intraclass correlations (ICC = 0.77 and 0.59, respectively) and low marginal R² values (0.09-0.11), indicating that individual differences explained most of the variance while group allocation contributed little. Additionally, the 25-min nap opportunity may not have been long enough to include deeper stages of sleep such as slow-wave sleep or rapid eye movement (REM) sleep, which are known to support memory consolidation, emotional regulation, and executive functioning (Genzel et al., 2014; Walker Matthew, 2009). As a result, the absence of these restorative stages could explain the lack of cognitive improvement following the nap condition. Similarly, NSDR, which does not involve actual sleep, is unlikely to activate the same neural mechanisms associated with cognitive enhancement. Together, these factors suggest that under well-rested conditions, the cognitive benefits of a short nap or NSDR are minimal and may not extend beyond perceptual or self-reported outcomes.

Handgrip strength showed a significant group × time interaction, but post-hoc comparisons revealed no significant differences between the nap, NSDR, and control conditions at any time point. This suggests that while performance patterns varied slightly over time, these changes were not significantly influenced by the type of intervention. Reliability and model fit indices support this interpretation. Handgrip strength demonstrated excellent reliability (ICC = 0.86), and the mixed model explained nearly all variance (conditional R² = 0.95), with a substantial proportion attributed to fixed effects (marginal R² = 0.63). These values indicate consistent measurement and limited potential for acute enhancements in strength across conditions. This outcome aligns with previous findings showing minimal effects of short daytime naps on muscle force production (Boukhris, Trabelsi, et al., 2024). Given the short duration and absence of prior fatigue, it is plausible that neither a 25-min nap nor an NSDR condition indues the neuromuscular processes necessary to enhance strength within such a brief post-intervention period. Sex was a significant covariate for handgrip strength, suggesting that observed variability was mainly attributable to sex-related differences in maximal force capacity rather than effects of the nap or NSDR.

The delayed effects observed after the nap may be explained by autonomic adjustments occurring during sleep. Immediately after the nap, greater amounts of light sleep and longer total sleep time likely contributed to higher sleepiness, higher fatigue, and lower readiness to perform, consistent with sleep inertia, which can last up to an hour after waking (Mesas et al., 2023). By 40 minutes post-nap, these responses had shifted in line with the performance outcomes. Reduced heart rate during the nap may have contributed to lower fatigue and greater readiness to perform once sleep inertia had dissipated. Previous studies have shown that naps, even of short duration, promote vagal dominance and support autonomic regulation, which in turn reduces perceived exertion and enhances readiness (Boukhris et al., 2023; Faraut et al., 2017). Taken together, these findings suggest that the benefits of short naps are less dependent on progression into deeper sleep stages and more closely tied to autonomic adjustments that restore physiological balance.

Although NSDR did not produce significant group-level improvements in perceptual or performance outcomes, autonomic modulation may still play a role in its effects. NSDR induces a wakeful state of deep relaxation, which is accompanied by enhanced vagal activity and reduced cardiovascular strain, mechanisms known to support attentional control, executive efficiency, and physiological regulation (Laborde et al., 2017; Shaffer & Ginsberg, 2017; Thayer et al., 2009). While no measurable effects were observed in the present study, NSDR may still facilitate a state of physiological relaxation that could be beneficial in applied settings.

A key distinction between the interventions likely lies in their underlying physiological mechanisms. The nap opportunity provided actual sleep, with effects emerging once sleep inertia had dissipated. These effects were accompanied by autonomic adjustments, reflected in lower heart rate and subsequent reductions in fatigue alongside greater readiness to perform. In contrast, NSDR induced a wakeful state of deep relaxation without transitioning into sleep. Its effects appeared to be mediated by autonomic modulation, specifically enhanced vagal activity and reduced cardiovascular load. This pattern suggests that naps exert their influence through sleep-dependent restorative processes, whereas NSDR operates through non-sleep mechanisms that promote parasympathetic activation and physiological calm. Accordingly, a nap may be more suitable when there is sufficient time for the dissipation of sleep inertia and the emergence of delayed benefits, while NSDR may offer a practical alternative for eliciting autonomic relaxation when actual sleep is not feasible.

Some limitations should be acknowledged. The absence of a true pre-intervention baseline makes it difficult to determine whether within-group changes reflect intervention effects or natural variability. However, a post-only design was intentionally adopted to minimise arousal, fatigue, and practice effects that could confound relaxation-based interventions such as NSDR and napping. While this approach reduces internal control, it offers a more ecologically valid representation of how these interventions would be implemented in applied settings. In addition, the lack of EEG data precluded a direct comparison of neural activity between nap and NSDR, limiting our ability to delineate mechanistic pathways with greater precision.

## Conclusion

A 25-min nap opportunity and a 10-min NSDR condition produced distinct patterns of response. The nap condition led to reductions in fatigue compared with the control condition and greater readiness to perform compared with both the control and NSDR conditions at 40-min post-intervention. These delayed effects likely reflect the dissipation of sleep inertia and the autonomic adjustments that accompany light sleep (N1 and N2), which together contribute to improved perceptual states. In contrast, NSDR did not produce significant changes across perceptual, cognitive, or physical measures. Overall, these results indicate that short naps can enhance perceived readiness and reduce fatigue when sufficient time is allowed post-awakening, whereas NSDR may still hold potential as a relaxation strategy but did not elicit measurable benefits under the conditions of this study.

## Ethics statement

The present study was approved by the Human Research Committee at La Trobe University.

## Data availability statement

The data that support the findings of this study are available from the corresponding author upon reasonable request.

