## Supplementary Material for "Comparing the effects of a short nap and non-sleep deep rest on perceptual, cognitive, and physical performance in active adults"

**Table S1.** Linear mixed-effects models results for perceptual, cognitive, and physical measures.

|  | Fixed effect | F | p | Effect size (ηₚ²) |
| --- | --- | --- | --- | --- |
| Sleepiness | Group | 0.36 | 0.69 | 0.01 |
|  | Time | 5.71 | 0.004 | 0.09 |
|  | Group × Time | 4.83 | 0.001 | 0.15 |
|  | Sex | 0.02 | 0.88 | 0.0004 |
|  | Sleep duration | 0.37 | 0.54 | 0.007 |
|  | Physical activity | 0.71 | 0.40 | 0.01 |
| Fatigue | Group | 0.53 | 0.59 | 0.02 |
|  | Time | 0.21 | 0.81 | 0.003 |
|  | Group × Time | 8.74 | < 0.001 | 0.24 |
|  | Sex | 0.73 | 0.39 | 0.01 |
|  | Sleep duration | 0.13 | 0.71 | 0.002 |
|  | Physical activity | 0.91 | 0.34 | 0.02 |
| Readiness to perform | Group | 3.54 | 0.03 | 0.12 |
|  | Time | 1.66 | 0.19 | 0.03 |
|  | Group × Time | 9.16 | < 0.001 | 0.25 |
|  | Sex | 1.17 | 0.28 | 0.02 |
|  | Sleep duration | 0.04 | 0.83 | 0.0008 |
|  | Physical activity | 0.01 | 0.90 | 0.0002 |
| Reaction time for correct trials during Simon task | Group | 0.60 | 0.55 | 0.02 |
|  | Time | 16.83 | < 0.001 | 0.21 |
|  | Group × Time | 2.27 | 0.06 | 0.08 |
|  | Sex | 0.07 | 0.79 | 0.003 |
|  | Sleep duration | 0.27 | 0.60 | 0.005 |
|  | Physical activity | 0.16 | 0.69 | 0.003 |
| Accuracy percentage during Simon task | Group | 0.52 | 0.59 | 0.02 |
|  | Time | 2.53 | 0.08 | 0.05 |
|  | Group × Time | 1.25 | 0.29 | 0.04 |
|  | Sex | 0.07 | 0.79 | 0.003 |
|  | Sleep duration | 0.34 | 0.55 | 0.006 |
|  | Physical activity | 3.35 | 0.07 | 0.06 |
| Handgrip strength | Group | 0.33 | 0.71 | 0.01 |
|  | Time | 1.62 | 0.20 | 0.03 |
|  | Group × Time | 4.00 | 0.004 | 0.13 |
|  | Sex | 77.89 | < 0.001 | 0.60 |
|  | Sleep quality | 0.15 | 0.69 | 0.002 |
|  | Physical activity | 3.80 | 0.05 | 0.07 |

**Table S2.** Intra-class correlation coefficients (ICC), marginal R², and conditional R² for perceptual, cognitive, and physical measures.

|  | Adjusted ICC | Conditional R^2^ | Marginal R^2^ |
| --- | --- | --- | --- |
| Sleepiness | 0.581 | 0.622 | 0.099 |
| Fatigue | 0.752 | 0.773 | 0.086 |
| Readiness to perform | 0.809 | 0.837 | 0.145 |
| Mean reaction time during Simon task | 0.773 | 0.792 | 0.085 |
| Accuracy during Simon task | 0.588 | 0.631 | 0.105 |
| Handgrip strength | 0.856 | 0.946 | 0.625 |
